# Clinical and imaging characteristics of Parkinson’s disease with negative alpha-synuclein seed amplification assay

**DOI:** 10.1101/2025.10.01.25337085

**Authors:** Sarah M. Brooker, Jacopo Pasquini, Seung Ho Choi, David-Erick Lafontant, Seyed-Mohammad Fereshtehnejad, Yashar Zeighami, Piergiorgio Grillo, Giulietta M. Riboldi, Houman Azizi, Roqaie Moqadam, Un Jung Kang, Kelly N.H. Nudelman, Andrew Siderowf, Caroline M. Tanner, Thomas F. Tropea, Tatiana Foroud, Lana M. Chahine, Brit Mollenhauer, Kalpana M. Merchant, Douglas Galasko, Christopher S. Coffey, Roseanne D. Dobkin, Ethan G. Brown, Roy N. Alcalay, Daniel Weintraub, Kenneth Marek, Tanya Simuni, Paulina Gonzalez Latapi, Nicola Pavese, Kathleen L. Poston, the Parkinson’s Progression Markers Initiative

## Abstract

**Background:** The CSF alpha-synuclein seed amplification assay (CSFasynSAA) detects alpha-synuclein aggregation in over 90% of individuals with sporadic PD (sPD). However, the clinical characteristics of sPD with negative CSFasynSAA remain undefined.

**Objectives:** Describe clinical and neuroimaging characteristics of CSFasynSAA negative sPD individuals in the Parkinson’s Progression Markers Initiative (PPMI).

**Methods:** We identified sPD PPMI participants with a negative CSFasynSAA (SAA negative, n=80) or positive CSFasynSAA (SAA positive, n=856) result at baseline. For comparative analysis between groups we used a reduced dataset (n=79 SAA negative and n=237 SAA positive) propensity-score matched on age, sex, and time since clinical diagnosis. Clinical parameters, DAT-SPECT, and MRI brain volumetrics were analyzed.

**Results:** The SAA negative and matched SAA positive groups had similar motor performance on the MDS-UPDRS-part III and similar cognitive performance on the MoCA at baseline. The proportion with severe hyposmia was 12% for SAA negative versus 73% for SAA positive (*p* < 0.001). Per PPMI enrollment criteria all participants were classified as having an abnormal DAT-SPECT. There were no significant differences in median quantitative DAT-SPECT measures between groups. The SAA negative group showed a higher degree of atrophy in subcortical brain regions including substantia nigra. Longitudinally, 14.3% of SAA negative participants had a change in diagnosis, versus 0.9% of SAA positive participants.

**Conclusions:** At baseline, SAA negative sPD PPMI participants have a substantially lower rate of hyposmia, but otherwise cannot be readily distinguished from SAA positive participants based on clinical characteristics. However, SAA negative participants have a greater degree of subcortical brain atrophy, and approximately 1 out of 6 SAA negative participants received a change in diagnosis.

## Introduction

Pathologically, Parkinson’s disease (PD) is characterized by intra-neuronal accumulation of alpha-synuclein (a-syn) within Lewy bodies and Lewy neurites. Until recently, confirmation of a-syn aggregation was only feasible via pathologic assessment of post-mortem brain tissue. As such, clinical trials use established clinical diagnostic criteria,^1^ and when enrolling newly diagnosed PD may supplement with a biomarker of presynaptic dopaminergic neuronal dysfunction such as dopamine transport (DAT)-SPECT.^2^ While DAT-SPECT increases diagnostic accuracy, DAT-SPECT abnormalities are non-specific for PD,^3^ and clinical trials using these criteria still may enroll a subset of individuals without Lewy-type pathology.

We have recently introduced a research framework for a biological definition of Neuronal Synuclein Disease (NSD) that includes biomarkers specific for identifying people with Lewy-type pathology.^4^ One use of this framework could be to increase the positive predictive value of the clinical diagnosis for presence of Lewy-type pathology early in the disease course when the clinical diagnosis may be less accurate. This framework leverages advances in a-syn biomarker development, specifically the a-syn seed amplification assay from cerebrospinal fluid (CSFasynSAA).^5^ CSFasynSAA has emerged as a reliable biomarker for detection of aggregated a-syn, and CSFasynSAA has been validated as a highly sensitive and specific technique for differentiating those with a clinical diagnosis of PD from healthy control individuals.^6–8^

Across multiple cohorts, including the Parkinson’s Progression Markers Initiative (PPMI), over 90% of people with sporadic PD (sPD) have positive CSFasynSAA.^9, 10^ Notably, amongst people with PD and pathogenic *LRRK2* variants, approximately one third have negative CSFasynSAA and an absence of Lewy-type pathology on post-mortem autopsy.^9, 11, 12^ Thus, genetic factors likely play a key role in regulating disease pathogenesis and biomarker results must thus be interpreted in the context of genetic results when available. However, for those individuals without *LRRK2* variants or other identified genetic etiologies for PD it remains unknown how those with clinical sPD with a negative CSFasynSAA (SAA negative), differ from those with positive CSFasynSAA (SAA positive). Answering this question could clarify the potential utility of the CSFasynSAA assay, and the application of the NSD framework, to clinical trial design. Specifically, if the progression of SAA negative individuals differs from those who are SAA positive, inclusion of SAA negative individuals into clinical trials could introduce heterogeneity that is unrelated to Lewy-type pathology.

To address this knowledge gap, we aimed to determine whether sPD participants in the PPMI cohort who are SAA negative at baseline have distinct clinical and neuroimaging characteristics compared to SAA positive participants. We also sought to evaluate longitudinal disease progression amongst SAA negative sPD participants and determine whether the diagnosis of PD in these individuals was revised over time.

## Methods

### Study population

Data were from PPMI, a multicenter prospective cohort study. The PPMI study protocol, including biofluid collection techniques and storage processes, have been published elsewhere in detail.^13^ Participants included in the study were those enrolled in the sPD PPMI cohort prior to March 10, 2025, who had CSFasynSAA performed at baseline enrollment. The inclusion criteria for enrollment in the sPD PPMI cohort were: 1) clinical diagnosis of PD, 2) time since clinical diagnosis of 2 years or less, and 3) abnormal DAT-SPECT based on study criteria which includes visual review by at least two nuclear medicine physicians with experience in reading DAT-SPECT imaging. The PPMI exclusion criteria for enrollment in the sPD PPMI cohort are detailed in full at ppmistudy.org. Of note, treatment with dopaminergic medication within 60 days of enrollment is one of the exclusion criteria. Participants were additionally excluded from this analysis if they had: 1) inconclusive or type II CSFasynSAA result at baseline analysis (see below for details), 2) were PPMI participants with a pathogenic variant in PD-associated genes (see below for details).

### CSFasynSAA

CSF analysis was performed at the baseline visit for all participants. Assessment of aggregated a-syn was performed via a CSFasynSAA completed by Amprion (Amprion Inc, San Diego, CA) as previously described.^9, 14^ There are two protocols that have been used in PPMI, the Amprion 150hr assay and the newer Amprion 24hr assay. The following assay parameters were used to define CSFasynSAA results: Fmax (highest fluorescence intensity), T50 (time to 50% of Fmax), and TTT (time to reach a target relative fluorescence unit threshold). Based on interpretation of these parameters, the following possible results were obtained: negative CSFasynSAA (SAA negative), positive CSFasynSAA (SAA positive), and inconclusive. The positive 24hr CSFasynSAA shows a distinct type I pattern in PD subjects and type II pattern in individuals with a pathologically confirmed diagnosis of multiple system atrophy (MSA).^14^ Therefore, for participants for whom both the 150hr and 24hr CSFasynSAA were performed, results of the 24hr CSFasynSAA were used to determine CSFasynSAA status. The number of SAA negative participants who had the 150hr and 24hr CSFasynSAA protocol and the concordance between assay results is detailed in Table S1. Regarding blinding, an elective return of research information (RORI) PPMI initiative was initiated in December 2023 so some participants and investigators may have been informed of CSFasynSAA results. Future work is needed to determine whether RORI has impacted investigator diagnosis classifications in PPMI.

### Clinical Assessments

Demographic data was collected per the PPMI protocol as previously reported, including age at enrollment, sex, race, Hispanic or Latino ethnicity, and education.^13^ Time since diagnosis at date of enrollment and family history of PD were recorded. The PPMI study includes clinical measures assessing motor performance, non-motor symptoms, and cognitive function conducted annually. This study included data on Hoehn & Yahr Stage, Movement Disorder Society-Unified Parkinson’s Disease Rating Scale (MDS-UPDRS)-Parts I, II, III, and IV scores, percentile score on the University of Pennsylvania Smell Identification Test (UPSIT), Modified Schwab & England Activities of Daily Living score, REM Sleep Behavior Disorder (RBD) screen questionnaire score (RBDSQ), Geriatric Depression Scale (GDS), and Scales for Outcomes in Parkinson’s disease-Autonomic (SCOPA-AUT), including total SCOPA-AUT score and subscores in each autonomic domain. For cognitive function, the Montreal Cognitive Assessment (MoCA) score was used as well as investigator diagnosis of cognitive status (normal, mild cognitive impairment, or dementia). Per PPMI protocol, at enrollment sPD participants are not allowed to be on dopaminergic medications. Once participants initiate dopaminergic therapy MDS-UPDRS Part III is assessed in medication OFF and ON states. The percentage of participants who initiated dopaminergic medications was assessed at 1-year follow-up. For those participants on dopaminergic medications at 1 year the difference between MDS-UPDRS Part III scores ON medication and OFF medication was calculated. To assess motor asymmetry, a motor symptom asymmetry index was calculated as follows: lateralized items on MDS-UPDRS-part III were assessed (items 3.3, 3.4, 3.5, 3.6, 3.7, 3.8, 3.15, 3.16, and 3.17) and the following calculation was made: absolute value of (right side – left side)/(right side + left side). At annual visits the PPMI site investigator makes a clinical determination of the most likely diagnosis. For this study Dementia with Lewy Bodies was not considered a change in diagnosis as it is a neuronal synuclein disease.

### Genetic testing

Genotyping methods for the PPMI study are described at ppmi-info.org. Participants with known pathogenic variants identified in the *LRRK2*, *GBA1*, *PRKN*, or *SNCA* genes were excluded from analysis. The following *LRRK2* pathogenic variants were tested for: G2019S, R1441C, R1441G, R1441H, I2020T, and N1437H. Genetic testing for the *LRRK2* G2019S variant was available for 76/80 (95%) of SAA negative sPD participants and was negative in all SAA negative sPD cases tested. Assessment for the complete set of pathogenic *LRRK2* variants was available for 58/80 (73%) of SAA negative participants and one individual in the SAA negative sPD group was found to have a *LRRK2* R1441G variant. The positive *LRRK2* genetic result in that SAA negative participant was received after primary analysis was completed and that participant was thus not excluded from the cohort.

### Dopamine transporter imaging analysis

DAT-SPECT at baseline visit was used to evaluate dopamine dysfunction in SAA negative and SAA positive groups. Dopamine dysfunction was assessed quantitatively by the following parameters: lowest putamen specific binding ratio (SBR), percent expected for age and sex, mean striatum binding, mean caudate binding, mean putamen binding, and the DAT binding asymmetry index, calculated as previously described.^15^

### Magnetic Resonance Imaging (MRI) analysis

MRI sequences for PPMI are described at ppmi-info.org (https://www.ppmi-info.org/sites/default/files/docs/PPMI2.0_002_MRI_TOM_Final_v4.0_20221118_FE.pdf). T1-weighted MRI at baseline visit was used to evaluate atrophy patterns between SAA negative and SAA positive sPD groups. Similar to our previous work, deformation-based morphometry (DBM) was used as the main measure of regional brain atrophy.^16–18^ Briefly, all T1-weighted images were pre-processed including denoising,^19^ non-uniformity correction,^20^ and intensity normalization. All images were then registered linearly and nonlinearly to MNI-ICBM-2009c template. All steps underwent visual quality control and participant scans that didn’t pass the quality control in any step were excluded from analysis (67/516 scans ~ 12.98%), consistent with the PPMI dataset failure rate at 13.10%. Voxel-wise DBM maps were generated by calculating the determinant of the Jacobian based on the deformation field based on the nonlinear transformations from subject T1-weighted to the template. These voxel-wise DBM maps were then averaged using an accurate atlas of 22 subcortical brain regions based on integration of BigBrain and MNI-ICBM-2009c template to calculate the regional DBM values for each participant scan for the statistical analysis.^21^ MRI analysis was carried out with the matched subset of subjects based on propensity-score matching as well as the complete unmatched sample. All neuroimaging results are reported as significant after False Discovery Rate (FDR) correction for multiple comparisons using a threshold of 0.05.

### Longitudinal outcomes

Disease progression was assessed using time to domain-based milestones as described.^22^ This methodology assesses 25 disease milestone metrics across six domains: “walking and balance,” “motor complications,” “cognition,” “autonomic dysfunction,” “functional dependence,” and “activities of daily living.” For each domain the participant was designated as having progressed to meet that domain if they had reached any of the previously defined milestone criteria within that domain. For example the “walking and balance” domain milestones are as follows: walking and balance impairment defined as MDS-UPDRS item 2.12 response ≥ 3; freezing of gait defined as MDS-UPDRS item 2.13 response ≥ 3 or MDS-UPDRS item 3.11 response = 4 (ON or OFF medications); gait impairment defined as MDS-UPDRS item 3.10 response ≥ 3 (ON or OFF medications); postural instability defined as MDS-UPDRS item 3.12 response ≥ 3 (ON or OFF medications); or Hoehn & Yahr stage ≥ 4. Details on milestone criteria for the other domains can be found as previously reported by Brumm et al.^22^ Other outcomes assessed include Hoehn & Yahr stage ≥ 3, moderate to severe non-dyskinesia motor complications defined as a score of ≥ 3 on MDS-UPDRS items 4.3 or 4.4, and research diagnosis.

### Neuropathologic assessment

Autopsy evaluation of post-mortem brain tissue was available for only one SAA negative sPD participant. Neuropathologic assessment was performed as described previously.^23^

### Statistical Analysis

Baseline demographic, clinical, and DAT-SPECT characteristics are summarized for SAA negative and SAA positive sPD groups, using median (interquartile range, IQR) and/or mean (standard deviation, SD) for continuous measures and frequency (percentage) for categorical measures. Differences in baseline characteristics between groups were assessed using the Wilcoxon rank sum test for continuous variables, and Chi-Square (or Fisher’s exact test when at least one expected cell count is below 5) for categorical variables.

For longitudinal survival analyses, Kaplan-Meier curves were generated for each endpoint as defined by Brumm et al.^22^: any progression milestone, 6 domain milestones; Hoehn & Yahr stage ≥ 3; MDS-UPDRS item 4.3 or 4.4 ≥ 3. These curves are presented by group over a 3-year period from enrollment, with at-risk tables and log-rank p-value. Participants who did not experience an event during the follow-up period were censored at the time of their last assessment within 3 years. Participants who had already reached any progression milestone or either of the two other survival outcomes at the baseline visit were excluded from analysis.

To account for age differences at enrollment between SAA negative and SAA positive sPD groups, a propensity-score matching approach was employed where SAA positive sPD participants were matched to the SAA negative sPD group based on age, sex, and time since diagnosis. The matching process utilized a caliper width of 0.25 times the standard deviation of the logit of the propensity score with an optimal fixed-ratio matching procedure designed to maximize the number of matched units while minimizing the standardized mean differences. One SAA negative participant was excluded from matched analysis due to missing data on time since clinical diagnosis.

Statistical analyses were performed using SAS v9.4 (SAS Institute Inc., Cary,NC, USA; sas.com; RRID:SCR_008567). Sankey diagrams were prepared using the “ggsankey” package in R (version 4.1.1; R Core Team 2024).

### Data Sharing

Data used in preparation of this article were obtained on March 10, 2025 from the PPMI database (www.ppmi-info.org/access-data-specimens/download-data), RRID:SCR_006431. For up-to-date information on PPMI, visit www.ppmi-info.org. This analysis was conducted by the PPMI Statistics Core and used actual dates of activity for participants, a restricted data element not available to public users of PPMI data. Statistical analysis codes used to perform the analyses in this article are shared on Zenodo (10.5281/zenodo.15238103).

## Results

### Sample Characteristics

Of the total sPD PPMI participants 80 were SAA negative compared to 856 who were SAA positive at baseline. Of the 80 SAA negative participants, repeat CSFasynSAA data at follow-up visits was available for 26. Twenty-two (84.6%) of these SAA negative participants remained SAA negative on follow-up while 4 (15.4%) had a type II result on follow-up. None of the SAA negative participants had a positive type I SAA on follow-up. Of the total 856 SAA positive sPD PPMI participants, repeat CSFasynSAA at a follow up visit was available for 309 participants. Of these, 292 (94.5%) of the SAA positive participants remained SAA positive at follow-up, 8 (2.6%) had a negative CSFasynSAA result, 5 (1.6%) had a type-II CSFasynSAA result, and 4 (1.3%) had an inconclusive CSFasynSAA.

### Baseline Demographic Characteristics of SAA negative sPD participants

The median age at enrollment for the SAA negative group was 66.8 years (IQR 61.7-73.1) compared to 63.9 years (IQR 57.2-69.9) in the SAA positive group, p=0.001 (Table S2). The median time since diagnosis and distribution of sex was comparable between groups (Table S2).

Due to the statistically significant between-group age difference, subsequent analyses used a reduced dataset matched for age, sex, and time since diagnosis, as described in the Methods section. The number of participants included in the matched dataset was n=79 SAA negative participants and n=237 SAA positive participants. Of note, one SAA negative participant was excluded from matched analysis due to missing data on time since clinical diagnosis.

### Baseline Clinical Characteristics

Baseline demographic and clinical characteristics of the matched SAA negative (n=79) and SAA positive (n=237) sPD groups are summarized in Table 1 and Table S3. Notably, the median percentile score on the UPSIT was 55.0 (IQR 26.0-78.5) for the SAA negative sPD group compared to 8.0 (IQR 4.0-16.0) for SAA positive. The percentage of participants with an UPSIT score less than or equal to the 15^th^ percentile was 12% for SAA negative versus 73% for SAA positive. The SAA negative group had higher scores on the MDS-UPDRS part II (p = 0.003), but there was no significant difference in MDS-UPDRS Part III scores in the OFF state (p = 0.648) (Fig. 1A). Analysis of tremor subscores demonstrated a lower rest tremor subscore in the SAA negative group compared to SAA positive (p = 0.002). Regarding non-motor features, there was no significant difference between groups in cognitive performance on the MoCA (p = 0.531) (Fig. 1A) or autonomic features as assessed by median SCOPA-AUT score (p=0.151).

**Figure 1:**
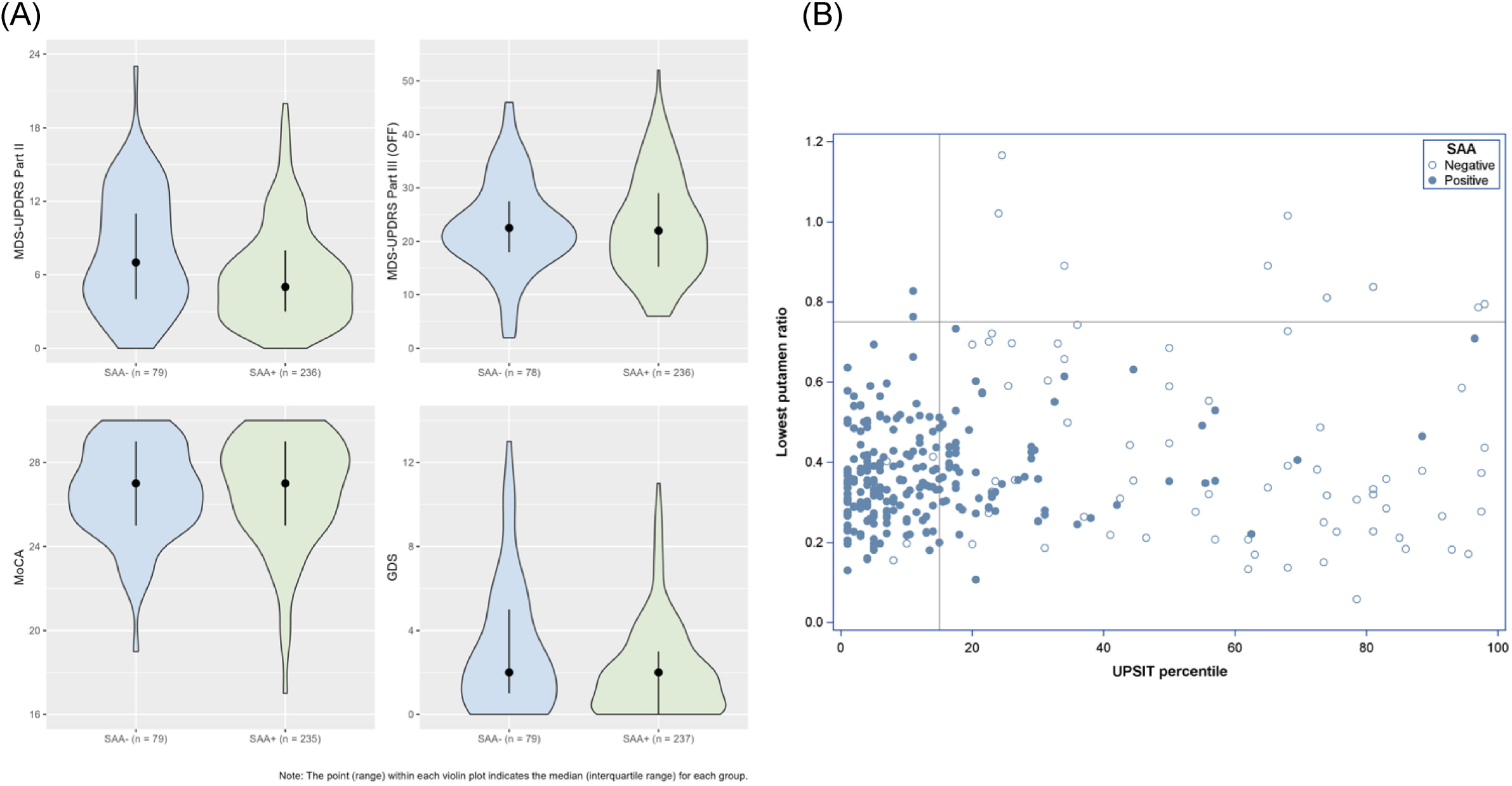
Baseline characteristics of SAA negative and SAA positive sPD participants. A) Violin plots of selected clinical characteristics measured at the baseline assessment are presented for SAA negative (SAA−) and SAA positive (SAA+) participants. The number of SAA− and SAA+ participants for whom data is available is indicated on the x-axis. For each plot the point indicates the median and the range indicates the interquartile range. (A) Median score on the MDS-UPDRS Part II, p = 0.003. (B) Median score on the MDS-UPDRS Part III in the off state, p = 0.648. (C) median score on the Montreal Cognitive Assessment (MOCA), p = 0.531. (D) median score on the geriatric depression scale (GDS), p = 0.025. B) The association between olfaction, dopamine transporter binding, and CSFasynSAA results is shown. Olfaction as indicated by percentile score on the UPSIT is shown on the x-axis and DATscan lowest putamen specific binding ratio is shown on the y-axis. SAA− participants are represented with an open circle and SAA+ participants are represented with a closed circle.

**Table 1:** Demographic and baseline characteristics of matched SAA− and SAA+ sporadic PD participants.

| Variable | SAA- sPD<br>(N = 79<sup>a</sup>) | SAA+ sPD<br>(N = 237) | p-value<sup>b</sup> |
| --- | --- | --- | --- |
| **Age at enrollment, years, median (IQR)** | 66.7 (61.3–73.3) | 67.4 (62.1–72.5) | 0.959 |
| **Male sex, n (%)** | 50 (63%) | 150 (63%) | 1.000 |
| **Family history of PD, n (%)** |  |  | 0.752 |
| 1st-degree family w/ PD | 10 (13%) | 37 (16%) |  |
| Non-1st-degree family w/ PD | 9 (11%) | 30 (13%) |  |
| No family w/ PD | 60 (76%) | 170 (72%) |  |
| **Time since diagnosis at enrollment, median (IQR)** | 0.5 (0.3–0.8) | 0.5 (0.3–0.8) | 0.369 |
| **Duration of follow-up from BL, years, median (IQR)** | 1.9 (1.0–3.0) | 2.1 (1.0–6.0) | 0.060 |
| **Hoehn & Yahr stage, n (%)** |  |  | 0.201<sup>c</sup> |
| 1 | 19 (24%) | 75 (32%) |  |
| 2 | 59 (75%) | 161 (68%) |  |
| 3 | 1 (1%) | 1 (<1%) |  |
| **UPSIT percentile, median (IQR)** | 55.0 (26.0–78.5) | 8.0 (4.0–16.0) | <.001 |
| **UPSIT ≤ 15th %ile, n (%)** | 9 (12%) | 171 (73%) | <.001 |
| **MDS-UPDRS Part I, median (IQR)** | 6.0 (4.0–11.0) | 5.0 (3.0–8.0) | 0.069 |
| **MDS-UPDRS Part II, median (IQR)** | 7.0 (4.0–11.0) | 5.0 (3.0–8.0) | 0.003 |
| **MDS-UPDRS Part III (OFF), median (IQR)** | 22.5 (18.0–27.0) | 22.0 (15.5–29.0) | 0.648 |
| **Motor symptom asymmetry index (OFF), median (IQR)** | 0.40 (0.20–0.60) | 0.52 (0.32–0.81) | 0.001 |
| **Tremor score (OFF), median (IQR)** | 4.0 (2.0–7.0) | 5.0 (3.0–7.0) | 0.075 |
| **Rest tremor subscore (OFF), median (IQR)** | 2.0 (0.0–4.0) | 4.0 (2.0–5.0) | 0.002 |
| **MDS-UPDRS Total Score (OFF), median (IQR)** | 39.0 (28.0–48.0) | 32.0 (25.0–43.0) | 0.020 |
| **MoCA, median (IQR)** | 27.0 (25.0–29.0) | 27.0 (25.0–29.0) | 0.531 |
| **Cognitive categorization, n (%)** |  |  | 0.244 |
| Normal | 54 (83%) | 150 (89%) |  |
| Mild cognitive impairment | 11 (17%) | 19 (11%) |  |
| **GDS, median (IQR)** | 2.0 (1.0–5.0) | 2.0 (0.0–3.0) | 0.025 |
| **SCOPA-AUT, median (IQR)** | 11.0 (7.0–14.0) | 9.0 (6.0–13.0) | 0.151 |
| **Lowest putamen ratio, median (IQR)** | 0.35 (0.23–0.60) | 0.35 (0.29–0.43) | 0.736 |
| **Mean striatum binding, median (IQR)** | 1.29 (1.03–1.83) | 1.40 (1.18–1.67) | 0.557 |
| **Mean caudate binding, median (IQR)** | 1.87 (1.45–2.33) | 1.93 (1.66–2.35) | 0.188 |
| **Mean putamen binding, median (IQR)** | 0.84 (0.56–1.39) | 0.84 (0.68–1.05) | 0.704 |
| **DAT binding asymmetry index, median (IQR)** | 0.12 (0.07–0.20) | 0.15 (0.08–0.25) | 0.169 |
| **Lowest putamen ratio ≥ 0.75, n (%)** | 9 (12%) | 2 (0.9%) | <.001 |

UPSIT = University of Pennsylvania Smell Identification Test; MDS-UPDRS = Movement Disorder Society-Unified Parkinson's Disease Rating Scale; MoCA = Montreal Cognitive Assessment; GDS = Geriatric Depression Scale; SCOPA-AUT = Scales for Outcomes in Parkinson's Disease-Autonomic
Missing data (if more than 5): Cognitive categorization $n = 82$ (consistent data collection on this variable began after 2020); DaTscan measures (lowest putamen ratio, mean binding, asymmetry index) $n = 6$ .
<sup>a</sup>One SAA- sPD participant excluded from matched analysis due to missing disease duration.
<sup>b</sup>Comparisons by SAA status used Chi-Square or Fisher's Exact tests for categorical variables and Wilcoxon rank sum tests for continuous variables.
<sup>c</sup>For the purposes of comparisons, Hoehn & Yahr stage was dichotomized as stage 1 vs. $\geq 2$ .

### Baseline DAT-SPECT metrics

All of the participants were classified as having an abnormal DAT-SPECT at baseline as per PPMI enrollment criteria, which included visual inspection with or without quantitative analysis. There were no significant differences between matched SAA negative and SAA positive sPD groups in the following quantitative DAT-SPECT imaging metrics at baseline assessment: median lowest putamen ratio, mean striatum binding, mean caudate binding, mean putamen binding, and the DAT binding asymmetry index (Table 1). However, while there was no difference between median lowest putamen striatal binding ratio between groups we found that 9/75 (12.0%) of the matched SAA negative participants had a lowest putamen ratio ≥ 0.75 compared to 2/235 (0.9%) of the matched SAA positive participants (p < .001). Four participants in the matched SAA negative group and 2 participants in the matched SAA positive group had missing lowest putamen ratio at baseline. The association between dopamine transporter binding, olfaction, and CSFasynSAA results is shown in Fig.1B.

### Baseline Structural MRI analysis

Compared to the propensity-score matched SAA positive group, SAA negative sPD participants had a significantly higher degree of atrophy in 9 out of 22 subcortical regions including bilateral substantia nigra, subthalamic nucleus, globus pallidus interna and externa, and left red nucleus (FDR corrected p-values < 0.05) (Fig. 2, Table S4). While not significant, the remaining 13 regions except for the right caudate showed a trend towards higher atrophy. To ensure the robustness of our findings, we repeated the analysis on the full unmatched sample, including Age and Sex as covariates. The results were consistent, except for the left putamen, which reached statistical significance in the full-sample analysis (Table S5).

**Figure 2:**
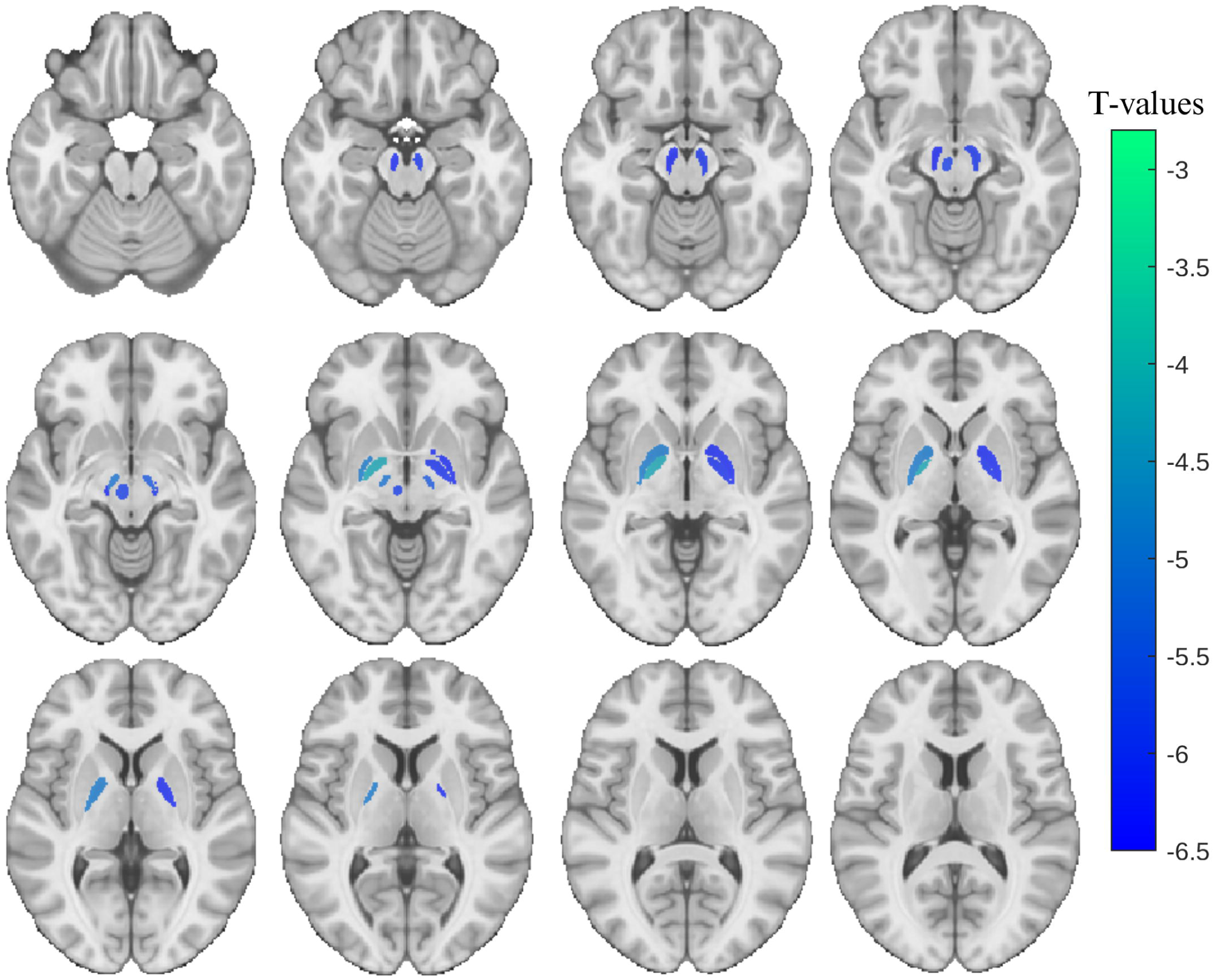
Cross-sectional brain atrophy map of the SAA negative sPD group. T-statistic maps (FDR-corrected p < 0.05) show regions where the SAA negative group exhibited significantly greater atrophy compared to the matched SAA positive group. Results are overlaid on axial slices of the MNI template, ranging from z = 58 (top left) to z = 91 (bottom right) in 3 mm increments. DBM: deformation based morphometry. The colormap indicates the t-values from the SAA negative group contrast between SAA negative and SAA positive participants from the linear model controlling for age and sex

### Longitudinal Analysis

Median follow-up from baseline was 1.9 years for the SAA negative group. At 2 years there was data available for 53% of SAA negative sPD participants. 39% of SAA negative participants had not yet reached the 2-year enrollment timepoint, 4% were lost to follow-up or were overdue for their 2-year visit, and 5% withdrew from the study (Table S6).

Compared to the propensity-score matched SAA positive sPD group, the SAA negative group had a faster time to reach any domain-based disease milestones (Fig. 3). The most notable difference in attainment of milestones was in the proportion of participants who met the walking and balance milestones, which includes complications such as freezing of gait and postural instability. The walking and balance milestones were met by 13% of SAA negative participants within 2 years compared to 3% of SAA positive. For those SAA negative participants who reached walking and balance milestones the most common milestone met was postural instability (Table S7).

**Figure 3:**
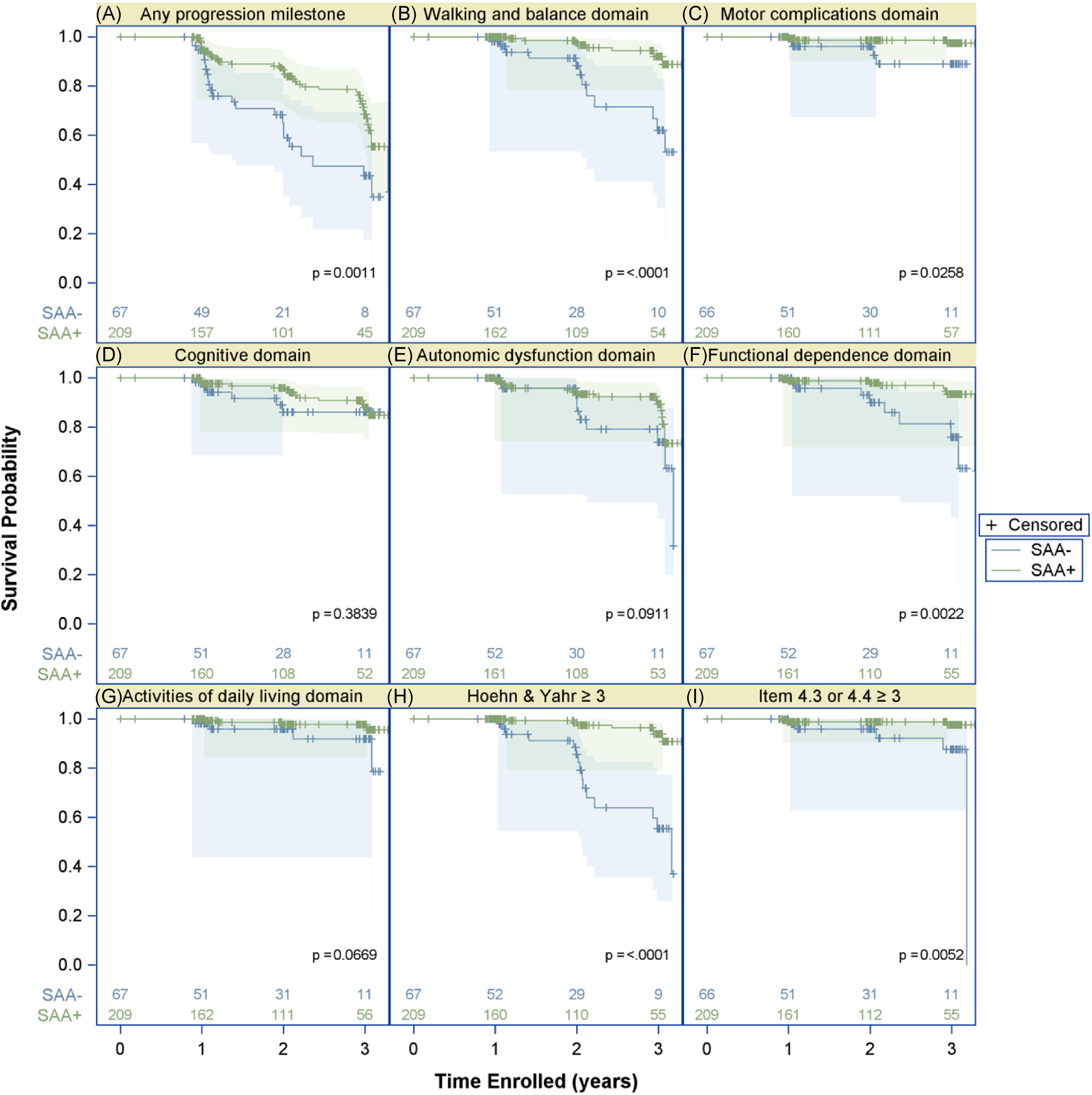
Time to reach disease progression milestones in matched SAA negative and SAA positive sPD. Kaplan-Meier curves are presented for time to reach disease progression milestones in the SAA negative (SAA−) and SAA positive (SAA+) matched groups over a 3-year time period. The number of SAA negative and SAA positive participants for whom data is available at each annual time point is indicated. The endpoints analyzed include: (A) Time to reach any of the 6 domain progression milestones defined by Brumm et al., (B) Walking and balance domain milestones, (C) Motor complications domain milestones, (D) cognitive domain milestones, (E) autonomic dysfunction domain milestones, (F) functional independence domain milestones, (G) activities of daily living domain milestones, (H) Hoehn & Yahr stage ≥ 3, and (I) moderate to severe non-dyskinesia motor complications defined as a score of ≥ 3 on the MDS-UPDRS item 4.3 or 4.4 which measure time in the off state and functional impact of motor fluctuations respectively.

### Dopaminergic treatment

At 1 year follow-up 33/66 (50%) of SAA negative participants and 126/202 (62.4%) of matched SAA positive participants had initiated dopaminergic treatment. Among the subset of matched participants who were on dopaminergic treatment, the median difference between MDS-UPDRS part III (ON) and MDS-UPDRS part III (OFF) at year 1 was not significant [−7.5 (−11, 0; n = 22) in the SAA negative group and −6.0 (−11, 0; n = 81) in the SAA positive group (p-value = 0.71)].

### Diagnostic change over time

Amongst the 80 SAA negative sPD participants, 77 had at least one follow-up visit and of these participants 11 (14.3%) had a change in research diagnosis at a follow-up visit (Fig. 4). By contrast, of the 237 matched SAA positive sPD group there were 231 who had at least one follow up visit, and only 2/231 (0.9%) had a change in diagnosis.

**Figure 4:**
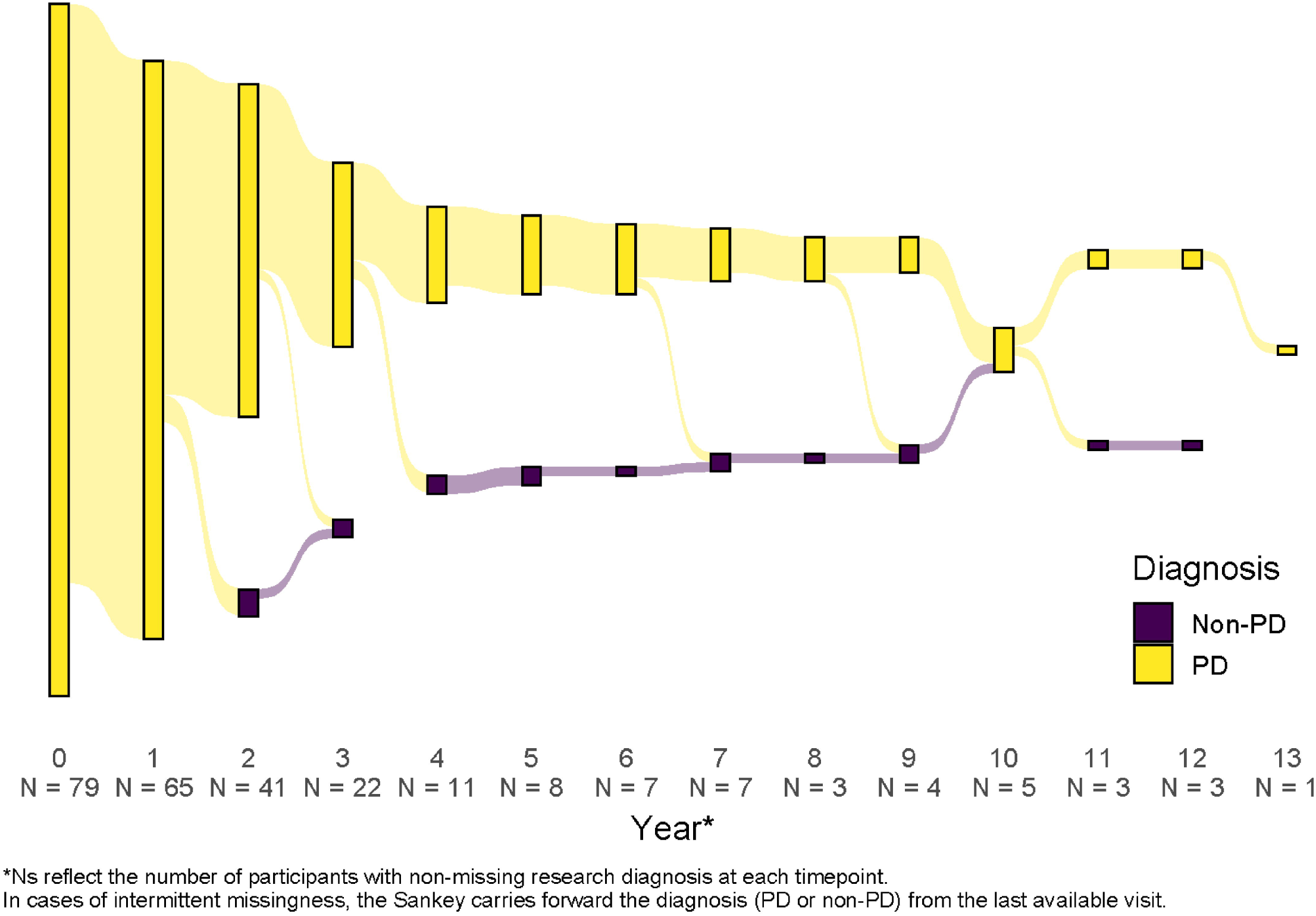
Sankey diagram of primary research diagnosis in SAA negative sporadic PD participants at annual visits. A sankey diagram is presented indicating the primary research diagnosis recorded at each annual follow-up visit for the SAA negative sPD group. Yellow indicates a diagnosis of Parkinson’s disease (PD) and purple indicates any other diagnosis that is not PD. The height of the bar at each year is proportional to the number of participants in each diagnosis category. For each annual timepoint the number of SAA negative participants for whom research diagnosis information was available is indicated on the x-axis.

In the SAA negative group, 6 participants (7.8%) had their research diagnosis changed to MSA ^13^. The other changed diagnoses were progressive supranuclear palsy (PSP), corticobasal syndrome (CBS), primary progressive freezing of gait, and prodromal synucleinopathy, each in one individual participant. In addition, one participant was deemed not to have PD or another neurologic disorder. Amongst the six participants for whom the diagnosis was changed to MSA, repeat CSFasynSAA data were available for two participants one of whom remained SAA negative and the other had a type II MSA-like result at 1-year follow-up. To assess whether elective return of research information (RORI) may have impacted investigator-determined diagnostic classifications, we evaluated the dates in which SAA negative participants accessed their CSFasynSAA results relative to the date of the change in diagnosis. We found that of the SAA negative participants who had a change in research diagnosis, none of these participants had accessed the CSFasynSAA results prior to the date of the change in diagnosis. To determine whether participants with a change in diagnosis impacted analysis of longitudinal disease progression we performed assessment of time to reach domain-based milestones with exclusion of the 11 SAA negative participants who had a change in research diagnosis. With these participants excluded, the SAA negative group still had a faster time to reach any domain-based disease milestone (p = 0.0023), and a faster time to walking and balance milestones (p = 0.0007) (Fig. S1).

### Pathologic Assessment

Post-mortem pathologic assessment of brain tissue was available for one of the SAA negative sPD participants. The research diagnosis for this individual was changed from PD to MSA at year 4 of follow-up. The participant died 7 years following enrollment in the study. Post-mortem brain tissue examination demonstrated glial cytoplasmic inclusions of a-syn consistent with a diagnosis of MSA-P.

## Discussion

In this study, we assessed clinical and neuroimaging characteristics of sPD participants in the PPMI cohort who lacked evidence of a-syn aggregation in the CSF and compared these participants to SAA positive sPD participants matched by key demographic characteristics. We found that there were several clinical features that differed between the two groups. Foremost amongst these was hyposmia with 73% of SAA positive participants performing at or less than the 15^th^ percentile on the UPSIT scale while only 12% of SAA negative sPD participants were hyposmic by this parameter. This finding is consistent with prior reports of high correlation between hyposmia and a-syn pathology.^9, 24^ However, the SAA negative and SAA positive sPD groups had similar baseline motor features and cognitive assessments and thus the data suggest that aside from hyposmia, standard clinical assessments alone at the time of clinical diagnosis are inadequate to identify individuals who may lack a-syn pathology. The strong concordance between hyposmia and CSFasynSAA positivity both in our analysis and in previous reports from the literature suggests that testing for hyposmia could be considered as a potential clinical marker for assessing the likelihood of a-syn pathology. Notably, although REM sleep behavior disorder is another common prodromal feature of synucleinopathies, we did not see a difference in median score on the REM sleep behavior disorder screening questionnaire between the SAA negative and SAA positive groups.

Using MRI deformation-based morphometry, the SAA negative participants showed significantly greater atrophy at a group level in multiple subcortical regions, including the substantia nigra, subthalamic nucleus, globus pallidus, putamen, and red nucleus. These differences were significant after matching for age, suggesting that the observed atrophy is not solely age-related. The underlying cause of the increased atrophy of subcortical brain regions in the SAA negative sPD group is unknown, but one hypothesis is that a subset of these individuals may in fact have atypical parkinsonian syndromes despite having a clinical diagnosis of sPD.

In our study, all individuals were classified as having a positive DAT-SPECT scan at baseline as this is part of the enrollment criteria into the sPD PPMI cohort. When evaluating the SAA negative and SAA positive sPD groups as a whole we found no differences between the SAA negative and SAA positive sPD groups in median quantitative DAT-SPECT metrics. Therefore SAA negative and SAA positive sPD individuals cannot be readily distinguished based on DAT-SPECT results alone. However, notably a subset of individuals had a lowest putamen ratio ≥ 0.75, which is used as a quantitative threshold for normal, despite having been classified as having a positive DATscan by visual interpretation and this proportion was higher in the SAA negative sPD group. This suggests that there can be discrepancies between visual interpretation of DAT-SPECT and quantitative metrics and that in clinical trial design detailed quantitative DAT-SPECT assessment in conjunction with fluid biomarker data could allow for more precise biological characterization of individuals with parkinsonism.

On longitudinal assessment, progression to clinically meaningful milestones appears to be faster in SAA negative individuals, but confirmation of the rate of disease progression in SAA negative sPD participants will require future longer longitudinal follow-up given the limitations of our small number of SAA negative participants with long-term follow up data. The SAA negative individuals had a notably higher rate of diagnostic change compared to SAA positive sPD individuals. This suggests that CSFasynSAA status could be important to consider in clinical trial design, particularly for a-syn targeting therapies. Notably, prior studies of the stability of the diagnosis of PD have shown high rates of change in diagnosis.^25–27^ But our finding that 14.3% of SAA negative sPD participants had a change in research diagnosis compared to only 0.9% of SAA positive sPD participants suggests that at least within the PPMI cohort CSFasynSAA status significantly impacts the stability of the PD diagnosis. This finding was not impacted by knowledge of CSFasynSAA status since we confirmed that those SAA negative individuals with a change in diagnosis had not accessed CSFasynSAA results prior to the research diagnosis change. It is important to stress that the results of our study should not be used to draw conclusions on the utility of CSFasynSAA in a clinical context in regards to making an initial diagnosis for individuals with parkinsonism, since by design our study was limited to individuals classified as sporadic PD in the PPMI research study, all of whom had DAT-SPECT imaging and met criteria for an sPD diagnosis. Thus, our cohort is not representative of the breadth of parkinsonian patients a clinician would encounter in the clinic.

When interpreting the study results, we consider potential causes of negative CSFasynSAA, including: 1) a true negative in an individual with a clinical syndrome indistinguishable from sPD, 2) a false negative in an individual with sPD and Lewy body pathology, and 3) a true negative in an individual who was erroneously diagnosed with sPD but actually has an alternative pathology, such as a 4-repeat tauopathy or glial cytoplasmic inclusions. To this last consideration, 14.3% of SAA negative sPD participants had a change in research diagnosis on follow-up. However, a significant proportion of the SAA negative sPD participants have not yet had extended longitudinal assessment so longer follow up will be required to determine if additional SAA negative participants receive a change in diagnosis over time. Given the possibility that some of the SAA negative sPD cohort may in fact have atypical parkinsonisms, comprehensive assessment of fluid biomarkers in the SAA negative sPD group is an important area for future studies, including assessment of neurofilament light chain in serum and CSF. Ultimately, post-mortem pathologic assessment will be required to determine the proportion of SAA negative participants who were mis-diagnosed as sPD and indeed have alternative neuropathologic diagnoses such as 4-repeat tauopathies or glial cytoplasmic inclusions, or vascular parkinsonism.

Regarding the possibility of false negative CSFasynSAA results, for the subset of SAA negative sPD participants for whom repeat CSF analysis was available on follow-up 85% remained SAA negative, 15% had a change to the type II MSA-like CSFasynSAA result and no participants had a change to type I CSFasynSAA positive. Therefore, the fairly high consistency of the results is evidence in support of a true negative result. The SAA positive sPD group also had a high rate of consistency in CSFasynSAA with 94.5% remaining SAA positive on follow-up. The one SAA negative participant with autopsy confirmation of glial cytoplasmic inclusions, and no Lewy body pathology, also supports this conclusion. However, additional neuropathological concordance data is necessary to interpret a SAA negative result in sPD as our data do not confirm that SAA negative status in parkinsonian individuals negates the possibility that they have Lewy-type pathology at autopsy. Therefore, we do not purport that this study should be extrapolated to the clinical setting and do not suggest that SAA negative parkinsonian patients be told they do not have a Lewy body disease based on a biomarker alone. Rather, clinicians should continue to use all of the clinical and biological data, including biomarkers when available, to make the best diagnosis.

The median age at disease onset for the SAA negative sPD group was significantly higher than the SAA positive group and we performed age-matching to account for the potentially confounding effects of age. However, the older age in the SAA negative group may in itself be biologically relevant as older individuals may be more likely to have non-Lewy body pathologies including 4-repeat tauopathies such as PSP or corticobasal degeneration.^28, 29^ In addition to identifying individuals with pathologic a-syn, there is an urgent need for development of biomarkers for other pathologies to improve diagnostic accuracy, assess co-pathology, and improve clinical trial design.

In this study, individuals with pathogenic variants in *LRRK2*, *GBA1*, *PRKN*, and *SNCA* were excluded as they were not classified as sporadic. Notably, amongst people with LRRK2-PD approximately one third lack a-syn containing Lewy body pathology in post-mortem brain tissue,^11, 12^ and 32.5% have negative CSFasynSAA results.^9^ Similarly, PD patients with pathogenic *PRKN* variants often lack neuronal a-syn on brain autopsy.^30, 31^ Since some genetic forms of PD lack pathologic a-syn, it is possible that a subset of the SAA negative participants in this study may have an as yet unrecognized genetic cause of PD resulting in a clinically indistinguishable, yet biologically distinct disease process.

### Limitations and future studies

A major limitation of this study is the limited long-term follow-up of SAA negative sPD participants. The PPMI study will continue longitudinal follow-up to characterize symptom progression and diagnostic change in the SAA negative sPD group. Another factor impacting interpretation of this study is the relatively small number of SAA negative sPD participants, which is reflective of the high rate of CSFasynSAA detection in sPD. One additional factor impacting study interpretation is that the SAA negative sPD group is heterogeneous with a subset of individuals likely falling into distinct disease categories, including those with MSA, PSP, or CBD, who were mis-diagnosed as sPD. An additional limitation is that while 76/80 (95%) of SAA negative participants were confirmed to be negative for the LRRK2 G2019S variant, complete genetic testing for the full list of *LRRK2* pathogenic variants was only available for 73% of SAA negative participants. One *LRRK2* positive SAA negative case was included in the cohort since the genetic result was received after primary analysis was already complete. But given that it was only a singular participant, the inclusion of this individual in our data set does not affect our results or conclusions. The rate of *LRRK2* positivity amongst individuals with PD is approximately 2.4%,^32^ and thus amongst the 22 SAA negative participants without complete genetic testing it is unlikely that any individuals have a pathogenic *LRRK2* variant. Even for those individuals for whom *LRRK2* testing was complete it is possible that some individuals could have a *LRRK2* variant of undetermined significance that has not been classified as pathogenic. We anticipate that at least a subset of SAA negative sPD cases may have genetic modifiers that could explain the lack of a-syn aggregation in CSF and thus future work is needed to perform detailed genetic analysis of the SAA negative sPD group to further investigate the influence of non-pathogenic genetic modifiers. Another limitation of the current study is the lack of inclusion of biomarker data regarding co-pathologies such as tau or more general biomarkers of neurodegeneration such as neurofilament light chain. This data is not yet available for the full cohort and thus future work will involve a comprehensive assessment of additional fluid biomarkers in the SAA negative sPD participants.

## Conclusion

SAA negative sPD PPMI participants have a substantially lower rate of hyposmia compared to SAA positive participants, but otherwise cannot be readily distinguished by baseline clinical characteristics. However, SAA negative participants have a higher degree of atrophy in subcortical brain regions and a higher proportion of SAA negative participants had a change in diagnosis compared to SAA positive participants. These data suggest that assessment of CSFasynSAA or alternative validated a-syn biomarkers should be considered in sPD clinical trials in which inclusion of individuals unlikely to have Lewy body pathology may confound trial data. Further longitudinal follow up of the SAA negative sPD PPMI participants will be critical to more fully assess disease progression over time in this group.

## Data Availability

Data used in preparation of this article were obtained on March 10, 2025 from the PPMI database (www.ppmi-info.org/access-data-specimens/download-data), RRID:SCR_006431. For up-to-date information on PPMI, visit www.ppmi-info.org

## Acknowledgements

PPMI – a public-private partnership – is funded by the Michael J. Fox Foundation for Parkinson’s Research and funding partners, including 4D Pharma, Abbvie, AcureX, Allergan, Amathus Therapeutics, Aligning Science Across Parkinson’s, AskBio, Avid Radiopharmaceuticals, BIAL, BioArctic, Biogen, Biohaven, BioLegend, BlueRock Therapeutics, Bristol-Myers Squibb, Calico Labs, Capsida Biotherapeutics, Celgene, Cerevel Therapeutics, Coave Therapeutics, DaCapo Brainscience, Denali, Edmond J. Safra Foundation, Eli Lilly, Gain Therapeutics, GE HealthCare, Genentech, GSK, Golub Capital, Handl Therapeutics, Insitro, Jazz Pharmaceuticals, Johnson & Johnson Innovative Medicine, Lundbeck, Merck, Meso Scale Discovery, Mission Therapeutics, Neurocrine Biosciences, Neuron23, Neuropore, Pfizer, Piramal, Prevail Therapeutics, Roche, Sanofi, Servier, Sun Pharma Advanced Research Company, Takeda, Teva, UCB, Vanqua Bio, Verily, Voyager Therapeutics, the Weston Family Foundation and Yumanity Therapeutics.

## Authors’ Roles

(1) Research Project: A. Conception, B. Organization, C. Execution; (2) Statistical Analysis: A. Design, B. Execution, C. Review and Critique; (3) Manuscript Preparation: A. Writing of the First Draft, B. Review and Critique.

S.M.B. 1A, 1B, 1C, 2A, 2C, 3A, 3B

J.P. 1A, 1B, 1C, 2A, 2C, 3B

S.H.C. 1B, 1C, 2A, 2B, 2C, 3B

D.E.L.: 1B, 1C, 2A, 2B, 2C, 3B

S.M.F. 1A, 1C, 2B, 2C, 3B

Y.Z. 1A, 1C, 2B, 2C, 3B

P.G. 1C, 2C, 3B

G.M.R. 1C, 2C, 3B

H.A. 1C, 2C, 3B

R.M. 1C, 2C, 3B

U.J.K. 1C, 2C, 3B

K.N.H.N. 1C, 2C, 3B

A.S. 1A, 2C, 3B

C.M.T. 1A, 2C, 3B

T.F.T. 1A, 2C, 3B

T.F. 1A, 2C, 3B

L.M.C. 1A, 1B 2C, 3B

B. M. 1A, 2C, 3B

K.M.M 1A, 2C, 3B

D.G. 1A, 2C, 3B

C.S.C. 1A, 2A, 2C, 3B

R.D.D. 1A, 2C, 3B

E.G.B. 1A, 2C, 3B

R.N.A. 1A, 2C, 3B

D.W. 1A, 2C, 3B

K.M. 1A, 1B, 2C, 3B

T.S. 1A, 1B, 2C, 3B

P.G.L 1A, 2C, 3B

N.P. 1A, 1B, 2A, 2C, 3B

K.L.P. 1A, 1B, 2A, 2C, 3B

## Financial Disclosures of all authors (for the preceding 12 months)

S.M.B receives funding from the NIH/NINDS and the Michael J. Fox Foundation. She has received travel expense reimbursement from the Parkinson Study Group and The Michael J Fox Foundation.

J.P. has no disclosures.

S.H.C, is funded by grants from the Michael J Fox Foundation for Parkinson’s Research.

D.E.L. is funded by grants from the Michael J Fox Foundation for Parkinson’s Research.

S.M.F. has no disclosures

Y.Z. has received research funding from the Fonds de recherche du Québec—Santé Chercheurs boursiers et chercheuses boursières en Intelligence artificielle, Natural Sciences and Engineering Research discovery grant, and Canadian Institutes of Health Research.

P.G. has no disclosures.

G.M.R. has no disclosures.

H.A. has received a PhD scholarship from Fonds de la Recherche du Québec—Santé and Parkinson’s Canada.

R.M. has no disclosures

U.J.K. is supported by research grants from NIH, Parekh Center for Interdisciplinary Neurology. U.JK. is on SAB for Amprion, Inc. and NurrON. He is a consultant for UCB, Lundbeck, and HanAll.

K.N.H.N. receives grant funding from the Michael J. Fox Foundation, the Alzheimer’s Association, the National Collegiate Athletic Association, the U.S. Department of Defense, and the NIH-NIA.

A.S. has been a consultant to the following companies in the past year: Acadia, Eli Lilly and Co, Neurocrine, Theravance, Cerevance, Spark/Roche, Boerhinger-Ingelheim, Wave Life Sciences, Inhibikase, Prevail, Mitzubishi and Alertity Therapeutics. He has served on DSMBs for the Huntington Study Group and The Healey ALS Consortium (Massachusetts General Hospital). He has received grant funding from the Michael J. Fox Foundation, NIA and NINDS.

C.M.T. declares consultancies for Neurocrine, Praxis (advisory board via the International Parkinson’s and Movement Disorder Society), Jazz/Cavion, Roche Genentech and Bial. CMT also declares grant support to her institution from The Michael J. Fox Foundation, National Institutes of Health, Gateway LLC, Department of Defense, Roche Genentech and Parkinson Foundation.

T.F.T. has received research support from NIH, The Michael J Fox Foundation, the Parkinson Foundation, and Eli Lilly and Company. Dr Tropea serves as a clinical trial advisory board member for Bial, has received travel expense reimbursement from the Parkinson Study Group, The Michael J Fox Foundation and the Parkinson Foundation, and speakers fees from Catalyst Medical Education.

T.F. receives funding from the NIH and Michael J. Fox Foundation and serves on several advisory committees for NIH funded studies (with < $1,000 each).

L.M.C. Financial disclosure: consulting fees and research support from the Michael J Fox Foundation. COI: none

B.M. has received honoraria for consultancy and/or educational presentations from GE, Bial, Roche, Biogen, AbbVie, Desitin and Amprion. BM is member of the executive steering committee of the Parkinson Progression Marker Initiative of the Michael J. Fox Foundation for Parkinson’s Research and has received research funding from Aligning Science Across Parkinson’s disease (ASAP, CRN), the German Parkinsonstiftung and the Michael J. Fox Foundation.COI: none

K.M.M.: Consultant for Axial Therapeutics, Asceneuron, Calico, Hanall, JnJ, Michael J. Fox Foundation, Nitrase Therapeutics, NuraBio, NRG Therapeutics, Rome Therapeutics, Schrodinger, Ventyx, private equity companies. Serves on the BoD for Envisagenics, Retromer Tx; serves on the SAB for Axial, Nitrase, NRG, Sinopia, Vanqua; Received research Funding support from Michael J. Fox Foundation and Honoraria from ASAP.

D.G. has received grant support from the NIH and Michael J Fox Foundation, and is a consultant for Eisai, Lilly, Artery Therapeutics and Cognition Therapeutics.

C.S.C. receives funding from the NIH/NINDS, MJFF, and PCORI.

R.D.D. receives research funding from the Michael J. Fox foundation and the VA office of rural health.

E.G.B. has received grant support for research from the Michael J. Fox Foundation, the National Institutes of Health, the Department of Defense, and Gateway Institute for Brain Research, Inc. He has received consulting fees from Guidepoint Inc.

R.N.A. research is funded by the Michael J. Fox Foundation, the Silverstein Foundation, the Parkinson’s Foundation and the Aufzien Family Center for the Prevention and Treatment of Parkinson’s Disease. He received consultation fees from Bexxion, Biogen, Biohaven, Capsida, Gain Therapeutics, Genzyme/Sanofi, Janssen, SK Biopharmaceuticals, Takeda and Vanqua Bio.

D.W. has received research funding or support from Michael J. Fox Foundation for Parkinson’s Research, International Parkinson and Movement Disorder Society (IPMDS), National Institute on Health (NIH) and the U.S. Department of Veterans Affairs; honoraria for consultancy from AbbVie, Boehringer Ingelheim, CHDI Foundation, Citrus Health Group, Negev Labs, Otsuka, Parkinson Study Group, Sage Therapeutics, Signant Health and Vanqua Bio; and license fee payments from the University of Pennsylvania for the QUIP, QUIP-RS and PDAQ.

K.M.:Consultant for Michael J Fox Foundation, CPP, Roche, Biohaven, Neuron23, Prothena, Lilly, Calico, ABli, Mitro, BMS, Novartis, Teva.

T.S. declares consultancies for AcureX, Adamas, AskBio, Amneal, Blue Rock Therapeutics, Critical Path for Parkinson’s Consortium, Denali, The Michael J. Fox Foundation, Neuroderm, Roche, Sanofi, Sinopia, Takeda, and Vanqua Bio; on advisory boards for AcureX, Adamas, AskBio, Biohaven, Denali, GAIN, Neuron23 and Roche; on scientific advisory boards for Koneksa, Neuroderm, Sanofi and UCB; and received research funding from Amneal, Biogen, Roche, Neuroderm, Sanofi, Prevail and UCB and an investigator for NINDS, MJFF, Parkinson’s Foundation.

P.G.L declares an early investigator award from The Michael J. Fox Foundation

N.P. has received consultancy honoraria form: Hoffmann-La Roche, Britannia, Bial, AbbVie, Teva; Speaker fees from: Bial, Britannia, AbbVie, GE Healthcare, Boston Scientific; Research Funding from Independent Research Fund Denmark, Danish Parkinson’s disease Association, Parkinson’s UK, Center of Excellence in Neurodegeneration (CoEN) network award, GE Healthcare Grant, Multiple System Atrophy Trust, Weston Brain Institute, EU Joint Program Neurodegenerative Disease Research (JPND), EU Horizon 2020 research, The Michael J. Fox Foundation, F. Hoffmann-La Roche, Medtronic, Symbyx.

K.L.P. is funded by grants from the NIH, Michael J Fox Foundation for Parkinson’s Research, the Knight Initiative for Brain Resilience, the Wu Tsai Neuroscience Institute, Lewy Body Dementia Association, Parkinson’s Foundation, American Parkinson’s Disease Association, and the Sue Berghoff LBD Research Fellowship. She has been on the Scientific Advisory Board for Amprion and has been a consultant for Novartis, Lilly, BioArctic, Biohaven, Curasen and Neuron23.

