## Supplementary material for "Clinical and imaging characteristics of Parkinson’s disease with negative alpha-synuclein seed amplification assay": Table S1

### **Supplementary Table 1. Summary of baseline CSFasynSAA results in participants with SAA- result on either assay.**

| **Result Pattern (150h Baseline ∕ 24h Baseline)** | **N** |
| --- | --- |
| **Included in SAA- cohort** | |
| 0 / 0 | 13 |
| 0 / X | 9* |
| 3 / 0 | 1 |
| X / 0 | 57 |
| **Excluded from SAA- cohort (due to prioritization of 24h assay)** | |
| 0 / 1 | 3 |
| 0 / 2 | 3 |

0 = Negative, 1 = Positive, 2 = MSA-like, 3 = inconclusive, X = no result available.

* Of the 9 participants for whom only the 150hr CSFasynSAA was available at baseline, 7 had the 24hr assay performed at a follow-up visit and the follow-up 24hr result was negative in all 7 cases.
