## Supplementary material for "Clinical and imaging characteristics of Parkinson’s disease with negative alpha-synuclein seed amplification assay": Table S2

### **Supplementary Table 2. Selected demographic and baseline characteristics of SAA- and SAA+ sporadic PD participants.**

| **Variable** | **SAA- sPD (N = 80)** | **SAA+ sPD (N = 856)** | ***p*-value^a^** |
| --- | --- | --- | --- |
| **Age at enrollment, years**, median (IQR) | 66.8 (61.7–73.1) | 63.9 (57.2–69.9) | 0.001 |
| Mean (SD) | 66.6 (9.1) | 63.2 (9.4) |  |
| **Male sex**, n (%) | 51 (64%) | 560 (65%) | 0.764 |
| **Disease duration at enrollment**, median (IQR) | 0.5 (0.3–0.8) | 0.5 (0.3–1.0) | 0.604 |
| Mean (SD) | 0.6 (0.5) | 0.7 (0.6) |  |
| Missing | 1 | 0 |  |

^a^ Comparisons by SAA status used Chi-Square or Fisher's Exact tests for categorical variables and Wilcoxon rank sum tests for continuous variables.
