## Supplementary material for "Clinical and imaging characteristics of Parkinson’s disease with negative alpha-synuclein seed amplification assay": Table S3

### **Supplementary Table 3. Extended demographic and baseline characteristics of matched SAA- and SAA+ sporadic PD participants.**

| **Variable** | **SAA- sPD (N = 79^a^)** | **SAA+ sPD (N = 237)** | ***p*-value^b^** |
| --- | --- | --- | --- |
| **Age at enrollment, years**, median (IQR) | 66.7 (61.3–73.3) | 67.4 (62.1–72.5) | 0.959 |
| Mean (SD) | 66.5 (9.2) | 66.6 (8.6) |  |
| **Male sex**, n (%) | 50 (63%) | 150 (63%) | 1.000 |
| **Education > 12 years**, n (%) | 65 (82%) | 206 (87%) | 0.266 |
| Missing | 0 | 1 |  |
| **Family history of PD**, n (%) |  |  | 0.752 |
| 1st-degree family w/ PD | 10 (13%) | 37 (16%) |  |
| Non-1st-degree family w/ PD | 9 (11%) | 30 (13%) |  |
| No family w/ PD | 60 (76%) | 170 (72%) |  |
| **Disease duration at enrollment**, median (IQR) | 0.5 (0.3–0.8) | 0.5 (0.3–0.8) | 0.369 |
| Mean (SD) | 0.6 (0.5) | 0.6 (0.5) |  |
| **Duration of follow-up from BL, years**, median (IQR) | 1.9 (1.0–3.0) | 2.1 (1.0–6.0) | 0.060 |
| **Hoehn & Yahr stage**, n (%) |  |  | 0.201^c^ |
| 1 | 19 (24%) | 75 (32%) |  |
| 2 | 59 (75%) | 161 (68%) |  |
| 3 | 1 (1%) | 1 (<1%) |  |
| **Race**, n (%) |  |  | 0.318^c^ |
| White | 75 (95%) | 215 (91%) |  |
| Black or African American | 4 (5%) | 6 (3%) |  |
| Asian | 0 | 4 (2%) |  |
| Other | 0 | 10 (4%) |  |
| Missing | 0 | 2 |  |
| **Hispanic or Latino ethnicity**, n (%) | 4 (5%) | 6 (3%) | 0.278 |
| Missing | 0 | 2 |  |
| **UPSIT percentile**, median (IQR) | 55.0 (26.0–78.5) | 8.0 (4.0–16.0) | <.001 |
| Missing | 1 | 4 |  |
| **UPSIT ≤ 15th %ile**, n (%) | 9 (12%) | 171 (73%) | <.001 |
| Missing | 1 | 4 |  |
| **MDS-UPDRS Part I**, median (IQR) | 6.0 (4.0–11.0) | 5.0 (3.0–8.0) | 0.069 |
| Missing | 1 | 2 |  |
| **MDS-UPDRS Part II**, median (IQR) | 7.0 (4.0–11.0) | 5.0 (3.0–8.0) | 0.003 |
| Missing | 0 | 1 |  |
| **MDS-UPDRS Part III (OFF)**, median (IQR) | 22.5 (18.0–27.0) | 22.0 (15.5–29.0) | 0.648 |
| Missing | 1 | 1 |  |
| **Motor symptom asymmetry index (OFF)**, median (IQR) | 0.40 (0.20–0.60) | 0.52 (0.32–0.81) | 0.001 |
| Missing | 0 | 1 |  |
| **Tremor score (OFF)**, median (IQR) | 4.0 (2.0–7.0) | 5.0 (3.0–7.0) | 0.075 |
| **Postural tremor subscore (OFF)**, median (IQR) | 1.0 (0.0–2.0) | 1.0 (0.0–1.0) | 0.324 |
| **Kinetic tremor subscore (OFF)**, median (IQR) | 1.0 (0.0–2.0) | 1.0 (0.0–2.0) | 0.671 |
| **Rest tremor subscore (OFF)**, median (IQR) | 2.0 (0.0–4.0) | 4.0 (2.0–5.0) | 0.002 |
| **MDS-UPDRS Total Score (OFF)**, median (IQR) | 39.0 (28.0–48.0) | 32.0 (25.0–43.0) | 0.020 |
| Missing | 2 | 3 |  |
| **Modified Schwab & England**, median (IQR) | 95.0 (90.0–100.0) | 95.0 (90.0–100.0) | 0.245 |
| **MoCA**, median (IQR) | 27.0 (25.0–29.0) | 27.0 (25.0–29.0) | 0.531 |
| Missing | 0 | 2 |  |
| **Cognitive categorization**, n (%) |  |  | 0.244 |
| Normal | 54 (83%) | 150 (89%) |  |
| Mild cognitive impairment | 11 (17%) | 19 (11%) |  |
| Missing | 14 | 68 |  |
| **RBDSQ**, median (IQR) | 3.0 (2.0–6.0) | 3.0 (2.0–5.0) | 0.912 |
| Missing | 0 | 4 |  |
| **pRBD (RBDSQ ≥ 6)**, n (%) | 20 (25%) | 51 (22%) | 0.530 |
| Missing | 0 | 4 |  |
| **GDS**, median (IQR) | 2.0 (1.0–5.0) | 2.0 (0.0–3.0) | 0.025 |
| **SCOPA-AUT**, median (IQR) | 11.0 (7.0–14.0) | 9.0 (6.0–13.0) | 0.151 |
| Missing | 2 | 3 |  |
| **SCOPA-AUT Gastrointestinal Score**, median (IQR) | 2.0 (1.0–4.0) | 2.0 (1.0–4.0) | 0.827 |
| Missing | 1 | 2 |  |
| **SCOPA-AUT Urinary Score**, median (IQR) | 5.0 (3.0–6.0) | 4.0 (3.0–6.0) | 0.534 |
| Missing | 1 | 2 |  |
| **SCOPA-AUT Cardiovascular Score**, median (IQR) | 0.0 (0.0–1.0) | 0.0 (0.0–1.0) | 0.277 |
| Missing | 1 | 1 |  |
| **SCOPA-AUT Thermoregulatory Score**, median (IQR) | 1.0 (0.0–2.0) | 1.0 (0.0–2.0) | 0.209 |
| Missing | 1 | 1 |  |
| **SCOPA-AUT Pupillomotor Score**, median (IQR) | 0.0 (0.0–1.0) | 0.0 (0.0–1.0) | 0.700 |
| Missing | 1 | 1 |  |
| **SCOPA-AUT Sexual Score**, median (IQR) | 0.0 (0.0–2.0) | 0.0 (0.0–2.0) | 0.581 |
| Missing | 2 | 1 |  |
| **Lowest putamen ratio**, median (IQR) | 0.35 (0.23–0.60) | 0.35 (0.29–0.43) | 0.736 |
| Missing | 4 | 2 |  |
| **Mean striatum binding**, median (IQR) | 1.29 (1.03–1.83) | 1.40 (1.18–1.67) | 0.557 |
| Missing | 4 | 2 |  |
| **Mean caudate binding**, median (IQR) | 1.87 (1.45–2.33) | 1.93 (1.66–2.35) | 0.188 |
| Missing | 4 | 2 |  |
| **Mean putamen binding**, median (IQR) | 0.84 (0.56–1.39) | 0.84 (0.68–1.05) | 0.704 |
| Missing | 4 | 2 |  |
| **DAT binding asymmetry index**, median (IQR) | 0.12 (0.07–0.20) | 0.15 (0.08–0.25) | 0.169 |
| Missing | 4 | 2 |  |

UPSIT = University of Pennsylvania Smell Identification Test; MDS-UPDRS = Movement Disorder Society-Unified Parkinson's Disease Rating Scale; MoCA = Montreal Cognitive Assessment; RBDSQ = REM Sleep Behavior Disorder-Screening Questionnaire; GDS = Geriatric Depression Scale; SCOPA-AUT = Scales for Outcomes in Parkinson's Disease-Autonomic

^a^One SAA- sPD participant excluded from matched analysis due to missing disease duration.

^b^Comparisons by SAA status used Chi-Square or Fisher's Exact tests for categorical variables and Wilcoxon rank sum tests for continuous variables.

^c^For the purposes of comparisons, Hoehn & Yahr stage was dichotomized as stage 1 vs. ≥ 2, and race was dichotomized as White vs. other.
