## Supplementary material for "Clinical and imaging characteristics of Parkinson’s disease with negative alpha-synuclein seed amplification assay": Table S4

### **Supplementary Table 4. Baseline MRI structural analysis from matched SAA- and SAA+ sPD participants**

| Region | Estimate | T-value | P-value |
| --- | --- | --- | --- |
| Left red nucleus | -0.7 | -3.44 | 0.000817497868509641 |
| Left substantia nigra | -0.64 | -3.03 | 0.00298901796475307 |
| Right substantia nigra | -0.7 | -3.26 | 0.00148094605841368 |
| Left subthalamic nucleus | -0.83 | -4.23 | 4.66304788499494E-05 |
| Right subthalamic nucleus | -0.72 | -3.81 | 0.000217608155873354 |
| Left globus pallidus externa | -0.83 | -4.36 | 2.72791419492869E-05 |
| Right globus pallidus externa | -0.63 | -3.07 | 0.00265822104432756 |
| Left globus pallidus interna | -0.98 | -5.1 | 1.28699784513741E-06 |
| Right globus pallidus Interna | -0.66 | -3.2 | 0.00175021191360466 |
