## Supplementary material for "Clinical and imaging characteristics of Parkinson’s disease with negative alpha-synuclein seed amplification assay": Table S5

### **Supplementary Table 5. Baseline MRI structural analysis from unmatched SAA- and SAA+ sPD participants**

| Region | Estimate | T-value | P-value |
| --- | --- | --- | --- |
| Left red nucleus | -0.53 | -3.07 | 0.00233992397240574 |
| Left substantia nigra | -0.54 | -2.83 | 0.00491841789504269 |
| Right substantia nigra | -0.58 | -3.02 | 0.0027473274785897 |
| Left subthalamic nucleus | -0.86 | -4.91 | 1.30793162711068E-06 |
| Right subthalamic nucleus | -0.73 | -4.28 | 2.39023871929471E-05 |
| Left putamem | -0.56 | -3.09 | 0.00215668355670961 |
| Left globus pallidus externa | -0.99 | -5.41 | 1.06916053191345E-07 |
| Right globus pallidus externa | -0.73 | -3.94 | 9.88996669618816E-05 |
| Left globus pallidus interna | -1.19 | -6.47 | 2.76862101826289E-10 |
| Right globus pallidus Interna | -0.77 | -4.16 | 3.94192207551866E-05 |
