## Supplementary material for "Clinical and imaging characteristics of Parkinson’s disease with negative alpha-synuclein seed amplification assay": Table S6

### **Supplementary Table 6. SAA- sporadic PD participant status at year 2.**

| **Status (at year 2)** | **All Participants (n = 80)** | **Pre-2020 Participants (n = 22)** | **Post-2020 Participants (n = 58)** |
| --- | --- | --- | --- |
| Completed^a^ | 42 (53%) | 20 (91%) | 22 (38%) |
| Not yet due | 31 (39%) | 0 | 31 (53%) |
| Lost to follow-up (1.0)^b^ *or* overdue (2.0) | 3 (4%) | 1 (5%) | 2 (3%) |
| Withdrew before reaching visit | 4 (5%) | 1 (5%) | 3 (5%) |

^a^19 participants withdrew/completed study after completing the following annual visit: year 2 (n = 4), year 3 (n = 5), year 4 (n = 2), year 5 (n = 2), year 6 (n = 1), year 7 (n = 2), year 9 (n = 1), year 10 (n = 2).

^b^1 participant did not complete year 2 visit and withdrew before completing subsequent visit.
