## Supplementary material for "Clinical and imaging characteristics of Parkinson’s disease with negative alpha-synuclein seed amplification assay": Table S7

### **Supplementary Table 7. Walking and balance milestones met by matched SAA- and SAA+ sporadic PD participants at first event within 2 years.**

| **Variable** | **SAA- sPD (N = 67)** | **SAA+ sPD (N = 209)** |
| --- | --- | --- |
| **Any walking and balance milestone** | **9 (13%)** | **6 (3%)** |
| Postural instability (item 3.12 ≥ 3 [ON or OFF]) | 6 (9%) | 3 (1%) |
| Walking and balance (item 2.12 ≥ 3) | 4 (6%) | 2 (1%) |
| Hoehn & Yahr (≥ 4 [ON or OFF]) | 2 (3%) | 1 (<1%) |
| Gait (item 3.10 ≥ 3 [ON or OFF]) | 2 (3%) | 2 (1%) |
| Freezing (item 2.13 ≥ 3) | 2 (3%) | 0 |
| Freezing of gait (item 3.11 = 4 [ON or OFF]) | 0 | 0 |

MDS-UPDRS = Movement Disorder Society-Unified Parkinson's Disease Rating Scale

Data only considers the *initial* event (i.e., first visit at which criteria for at least one walking and balance milestone were met). Columns include participants who did not meet any of the survival outcomes at baseline (i.e., excludes 12 SAA- and 28 SAA+ participants who met any of the survival outcomes at baseline, of whom 3 and 2 participants met a walking and balance milestone, respectively).
