## Supplementary material for "Clinical and imaging characteristics of Parkinson’s disease with negative alpha-synuclein seed amplification assay": Fig. S1

### **Supplementary Figure 1. Time to progression milestones in matched SAA- and SAA+ sporadic PD participants excluding SAA- participants who had change in primary research diagnosis**

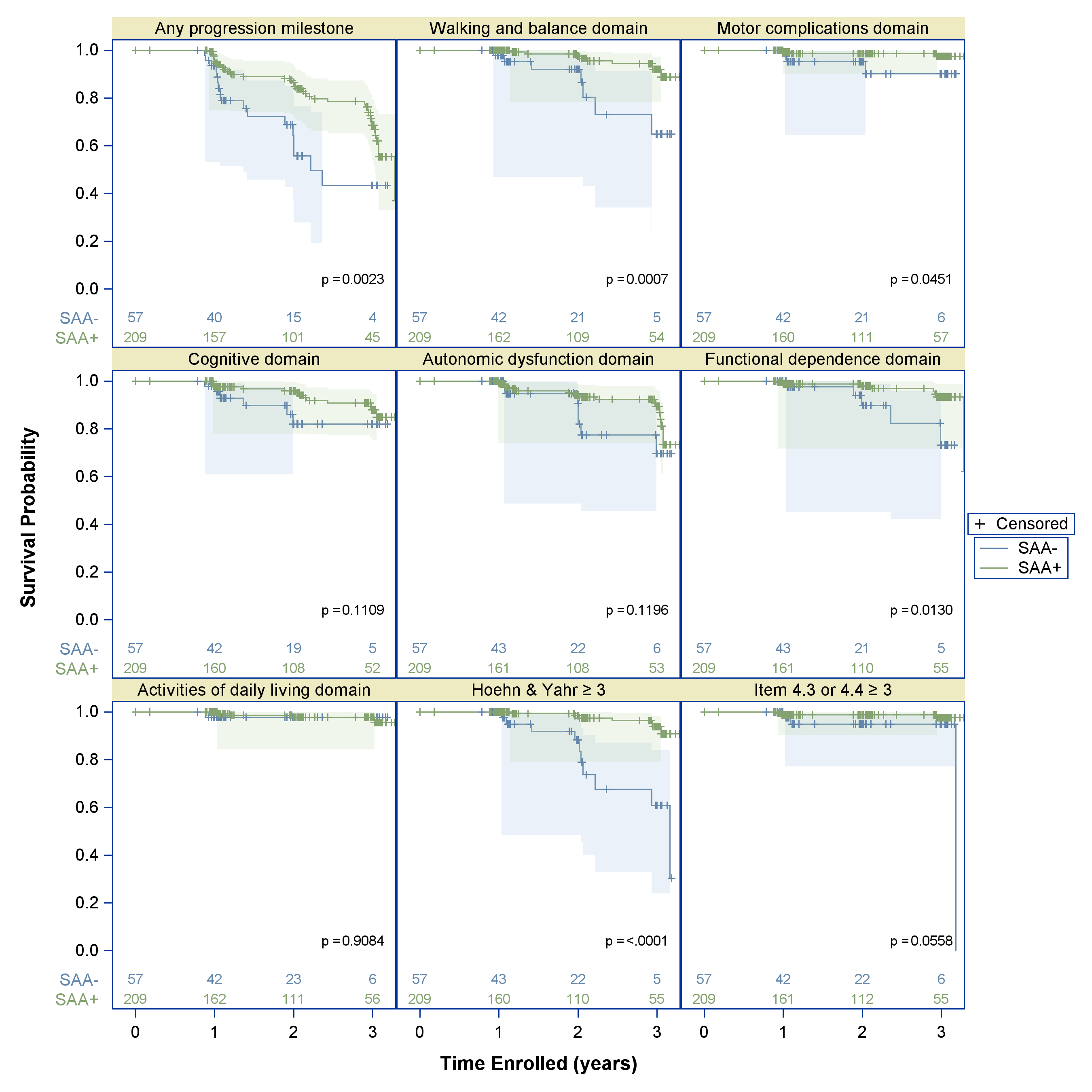
